# Comparing Existing Algorithms for Retrieving Pregnancy-related Adverse Event Reports

**DOI:** 10.64898/2026.02.17.26346457

**Authors:** Sara Hedfors Vidlin, Valentina Giunchi, Levente K-Pápai, Lovisa Sandberg, Cosimo Zaccaria, Takamasa Sakai, Loris Piccolo, Elena Rocca, Michele Fusaroli, Nhung TH Trinh

## Abstract

**Background:** Post-marketing surveillance is essential for complementing the safety profiles of medicinal products, especially for populations generally excluded from clinical trials such as pregnant individuals. However, the absence of a standardised pregnancy indicator in the electronic transmissions of adverse event reports hampers their correct identification in pharmacovigilance databases and complicates the study of safety concerns related to pregnancy exposures. Three recently developed rule-based algorithms with the common aim to systematically retrieve pregnancy-related reports differ in scope and are tailored to different databases (A. FAERS, B. EudraVigilance, C. VigiBase).

**Aim:** To compare the design and outputs of the three pregnancy algorithms.

**Methods:** This study was a collaboration among the authors of the three pregnancy algorithms. We harmonised their rules, implemented them in an R package to enable execution in both VigiBase and FAERS, and analysed key characteristics of reports flagged by each algorithm.

**Results:** The pregnancy algorithms A, B, and C flagged 235653, 279515, and 446957 reports respectively in VigiBase, and 265015, 260734, 350479 in FAERS. Reports exclusively retrieved by each algorithm (994, 3248, and 142324 in VigiBase, and 1528, 1100, and 59643 in FAERS) were mostly explained by Algorithm A having no age restriction, Algorithm B excluding normal pregnancy and ineffective contraception, and Algorithm C excluding paternal exposure.

**Conclusions:** Differences in flagging were largely related to varying scopes. Understanding commonalities and differences is crucial for empowering professionals working with pregnancy-related pharmacovigilance to select and use the most appropriate algorithm for their specific needs.

**Key points:**

- Three independently developed algorithms were designed to retrieve pregnancy-related adverse event reports and support research into pregnancy-specific safety concerns.
- By applying these algorithms to VigiBase and FAERS, we highlighted overlaps and differences in the reports they flag, reflecting heterogeneous scope and implementation.
- Awareness of these distinctions is essential to select and apply the most suitable algorithm for their specific needs.

## 1. Introduction

Pharmacovigilance refers to the science and activities relating to the detection, assessment, understanding and prevention of adverse effects or any other drug-related problem [1]. Clinical trials usually exclude pregnant individuals citing ethical considerations, resulting in limited safety data for this population at the time of marketing authorisation [2]. Post-marketing surveillance is therefore essential to assess potential risk during pregnancy. Adverse event reports, registries, and patient health records are useful for capturing safety information as medicinal products are used in broader, more diverse populations [3].

Adverse event reports are an important source to study safety concerns in pregnant individuals [4]. However, retrieving relevant reports is complicated due to the lack of a standardised indicator for flagging pregnancy cases in the current format for electronic transmission (E2B, issued by the International Conference on Harmonisation – ICH). To address this challenge, researchers have employed various strategies, such as searching among reactions and/or indications for terms included in the Standardised Medical Dictionary for Regulatory Activities (MedDRA®)^1^ Query (SMQ) titled ‘Pregnancy and neonatal topics’ or in similar manually customised queries [5–11]. These practical approaches may fall short in terms of performance.

Recently, three research groups have developed rule-based algorithms accounting for several structured data-elements to systematically flag pregnancy-related reports, aiming for better performance. Although these algorithms share a common goal, they were designed for different databases and reflect different choices in terms of scope. All three algorithms intend to retrieve reports concerning the pregnant person or the foetus/prenatally exposed child (without distinguishing between the two), and none includes exposure through lactation in the scope.

A. Sakai et al (2016) aims to flag pregnancy-related exposures in the Japanese Adverse Drug Event Reporting database (JADER) [12] and, in a later implementation, in the US Food and Drug Administration (FDA) Adverse Event Reporting System database (FAERS)^2^ [13]. The most recent implementation (available through the DiAna R package [14]) aimed for three different levels of accuracy; high sensitivity (i.e. recall), medium, and high positive predictive value (PPV; i.e. precision).
B. Zaccaria et al (2024) aims to flag pregnancy-related exposures, including paternal, and to exclude reports relating to menstrual disorders or contraception, or with no suspected adverse drug reaction, in EudraVigilance, the European database of suspected adverse reactions [15]. This algorithm achieved a PPV of 90% as measured on 100 randomly retrieved reports.
C. The VigiBase pregnancy algorithm described in Sandberg et al (2025) aims to flag pregnancy-related exposures, excluding paternal and regardless of occurrence of any adverse event, in VigiBase, the World Health Organization (WHO) global database of adverse event reports for medicines and vaccines [16]. The algorithm achieved a sensitivity of 75% (91% when restricting to the ICH-E2B reporting format) based on a dataset of 7874 reports, and a PPV of 92% based on 344 randomly retrieved reports.

Algorithm B is the only that explicitly formulates intended application, namely, signal detection. Although Algorithms B and C were evaluated in the database for which they were originally developed, their performance in other databases remains unknown. Moreover, differences between the algorithms have not been systematically compared. Understanding these aspects is important to make informed choices when exploring pregnancy-related safety issues in adverse event databases, for example in signal detection, descriptive studies, or addressing confounders.

## 2. Aim

We aimed to implement the three recently developed pregnancy algorithms, compare their foundational principles, and explore the characteristics of the reports they retrieve in FAERS and VigiBase.

## 3. Methods

### 3.1 Data sources

VigiBase, maintained by Uppsala Monitoring Centre (UMC), includes adverse event reports submitted by national pharmacovigilance centres in 160 countries as members of the WHO programme for international drug monitoring. The database structure is based on the ICH-E2B(R2) format and submitted reports are converted into this format. Reported adverse events and indications are mapped to the MedDRA terminology and medicinal products to the UMC’s Drug Dictionary WHODrug Global [17], a global medicine and vaccine terminology for identification of medicinal products. Data were retrieved until 2024-12-31, excluding suspected duplicates (n = 819368), as identified by vigiMatch [18], resulting in a dataset of 39.9 million reports.

FAERS, coordinated by the US FDA, contains adverse event reports from manufacturers as required by regulations (if serious and unexpected also from outside the US) along with reports received directly from consumers and healthcare professionals. Reports adhere to the ICH-E2B format, with adverse events coded to MedDRA Preferred Terms (PTs) and medicinal products left as free text. FAERS data files containing quarterly raw data of reports are made available to the public in both ASCII and XML format [19]. We used the processed version available via the DiAna R package [14], covering data up to 24Q4 (2024-12-31), where medicinal products are coded to the DiAna dictionary [20], resulting in a dataset of 18.6 million non-deduplicated reports.^3^

Reports in FAERS and VigiBase partially overlap, as domestic FAERS reports (originating from the US) are also included in the VigiBase dataset. Non-domestic reports are not included in the VigiBase dataset, as they are expected to be shared by the originating countries. Additionally, the VigiBase dataset was subjected to de-duplication, unlike the public version of FAERS.

### 3.2 Algorithms

We compared the algorithms from Sakai et al (high PPV version as Algorithm A), Zaccaria et al (Algorithm B), and Sandberg et al (Algorithm C). Supplementary analyses were conducted for the other versions of Sakai et al.

To enable comparison of the algorithms, we harmonised their rules for each data element. The implementation of the three algorithms and the script for the comparison analysis are made available through the open-source R package PVgravID, designed to be fully interoperable with the DiAna R package. Relevant data fields and list of terms utilised by the algorithms are described in Table S1 of the Electronic Supplementary Material (ESM). Authors’ original implementation flowcharts are available in Fig. S2:1-3 of the ESM.

### 3.3 Statistical and Graphical Comparison

We used Venn diagrams with proportional areas to illustrate the overlap and different flagging of reports by the three algorithms in FAERS and VigiBase.

Descriptive analyses were presented for demographic, clinical, and reporting variables, summarising categorical variables as absolute and relative frequencies, and continuous variables as median, interquartile range, and minimum and maximum values.

Upset plots were used to visualise the rules defining reports retrieved by: (1) only one algorithm, (2) two algorithms but not the third, and (3) all three algorithms.

## 4. Results

### 4.1 Implementation of the algorithms

**Fig. 1** illustrates the harmonised rule-out and rule-in steps across the three algorithms, highlighting their commonalities and differences. Algorithm A applies fewer but highly specific rule-in criteria, focusing on the route of administration and adverse events or indications associated with pregnancy exposure MedDRA PTs. Algorithm B and C share many logical and structural similarities: they first exclude reports based on rule-out criteria and then apply rule-in criteria to the remaining reports, although details in the specific rules differ. Compared to Algorithm A, they incorporate additional report features, such as gestation period and seriousness criterion, and use a broader range of pregnancy-related PTs beyond exposure terms, mainly based on sub-SMQs of the ‘Pregnancy and neonatal topics’ SMQ. None of the algorithms consider free text fields.

**Fig. 1.**
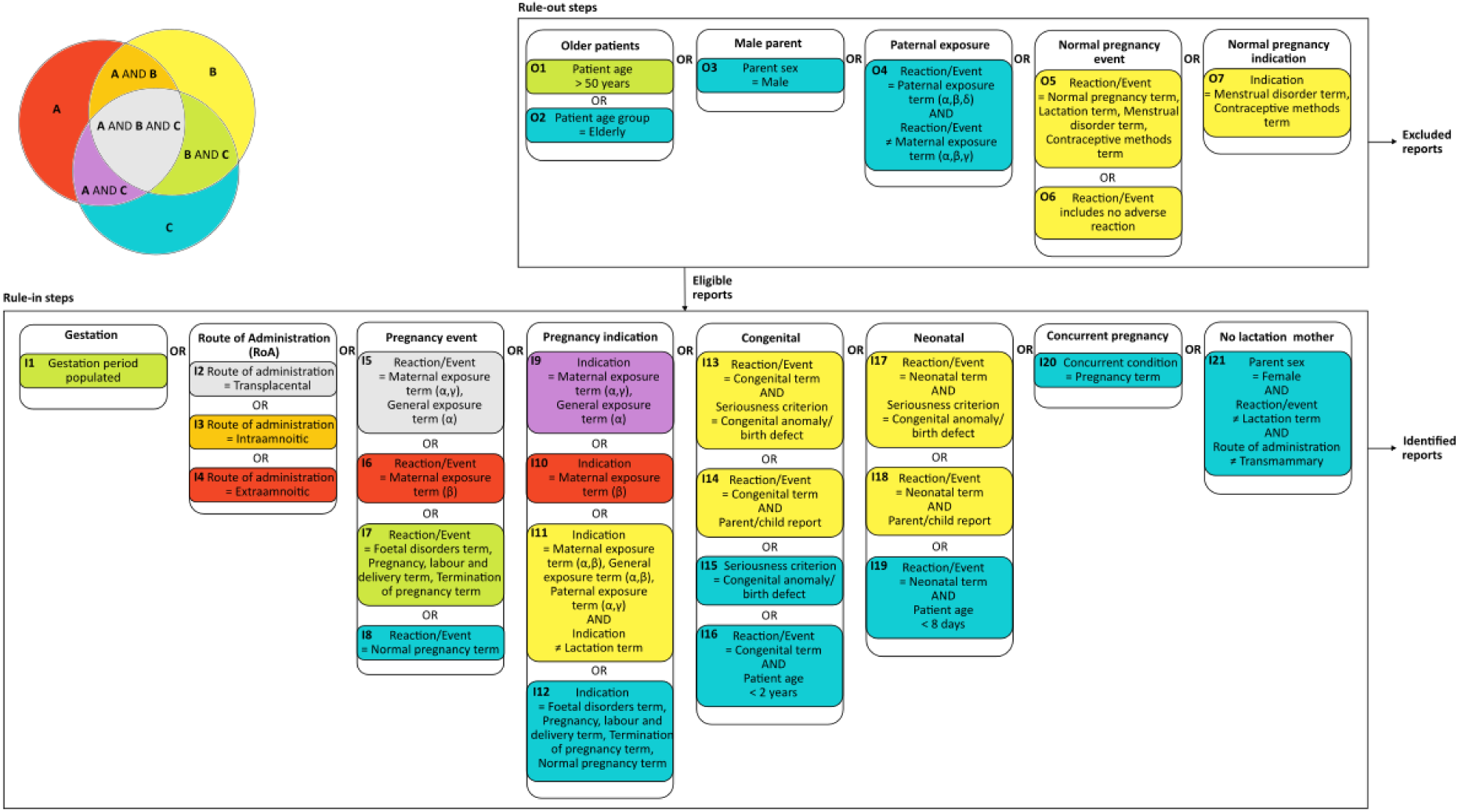
Implementation flowchart showing the rules of the algorithms, colour coded to highlight commonalities and differences. Terms in I5 are a subset of terms in I7, similarly, terms in I9 are a subset of terms in I12, these are broken out to visualise similarities. Terms in I7-I8, I12-I14, I16-I19 are based on sub-SMQs of the ‘Pregnancy and neonatal topics’ SMQ. Terms in I11 are all the non-lactation exposure terms in the MedDRA High Level Term (HLT) ‘Exposures associated with pregnancy, delivery and lactation’. See Table S1 of the ESM for complete list of terms for all criteria.

In VigiBase, 451703 reports were flagged as pregnancy-related by at least one algorithm. Of these, 205285 reports (45%) were retrieved by all three algorithms. Algorithm A flagged 994 reports (0.2%) not retrieved by any other algorithm, Algorithm B 3248 (0.7%), and Algorithm C 142324 (32%).

In FAERS, 353491 reports were flagged by at least one algorithm. Of these, 231517 reports (65%) were retrieved by all three. Algorithm A flagged 1528 reports (0.4%) not retrieved by any other, Algorithm B 1100 (0.3%), and Algorithm C 59643 (17%).

**Fig. 2** shows the overlap in flagged reports across the algorithms. Additional results for the other versions of the algorithm from Sakai et al are presented in Fig. S3:1-2 of the ESM.

**Fig. 2.**
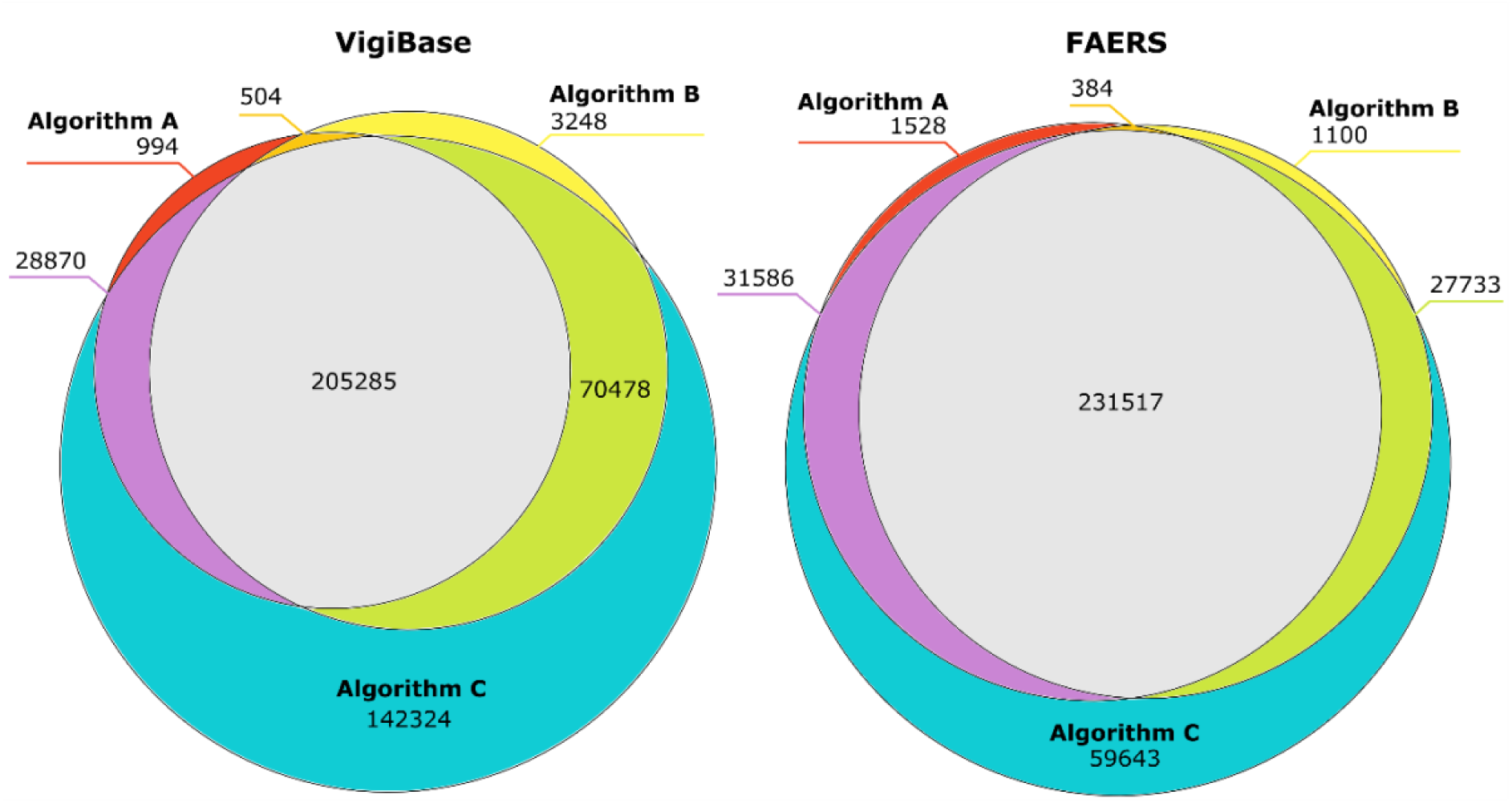
Venn diagrams showing the overlap of the pregnancy-related reports retrieved by the different algorithms, in VigiBase and FAERS, respectively.

### 4.2 Descriptive statistics of flagged pregnancy-related reports

All three algorithms flagged mostly reports concerning female patients in both databases (81-89%), see **Table 1**. Consumer reports constituted approximately one third of flagged reports in VigiBase, and half in FAERS. The age distribution of pregnancy-related reports was bimodal, with a smaller peak for neonates (7-13% in VigiBase, 15-21% in FAERS) and a bigger peak between 18-44 years (70-84%). Algorithm C retrieved a higher proportion of reproductive-age females and a smaller proportion of neonates (even if the absolute number was higher). Algorithm A was the only one that retrieved reports concerning patients older than 50 years (0.6-1%). Approximately half of the reports originated from the US and Canada, and one third from Europe, across algorithms and databases. Limited to VigiBase, Algorithms B and C retrieved more reports from Asia compared to Algorithm A (13% and 17%, respectively, vs 6%). Across all algorithms, the median number of adverse events per report was between two and three. Reports listing only one medication constituted 60-65% of the reports in VigiBase and 35-37% in FAERS, and 5-7% of VigiBase reports included more than five medications, compared to 20-22% in FAERS. The proportion of serious cases was 54-64% in VigiBase and 82-86% in FAERS. Hospitalisation and congenital anomaly were reported in 13-14% and 9-14% of all the retrieved reports in VigiBase, and in 18% and 17-21% in FAERS. Death was reported as a seriousness criterion in 2-3% of VigiBase reports and 5% of FAERS reports, across algorithms.

**Table 1.**
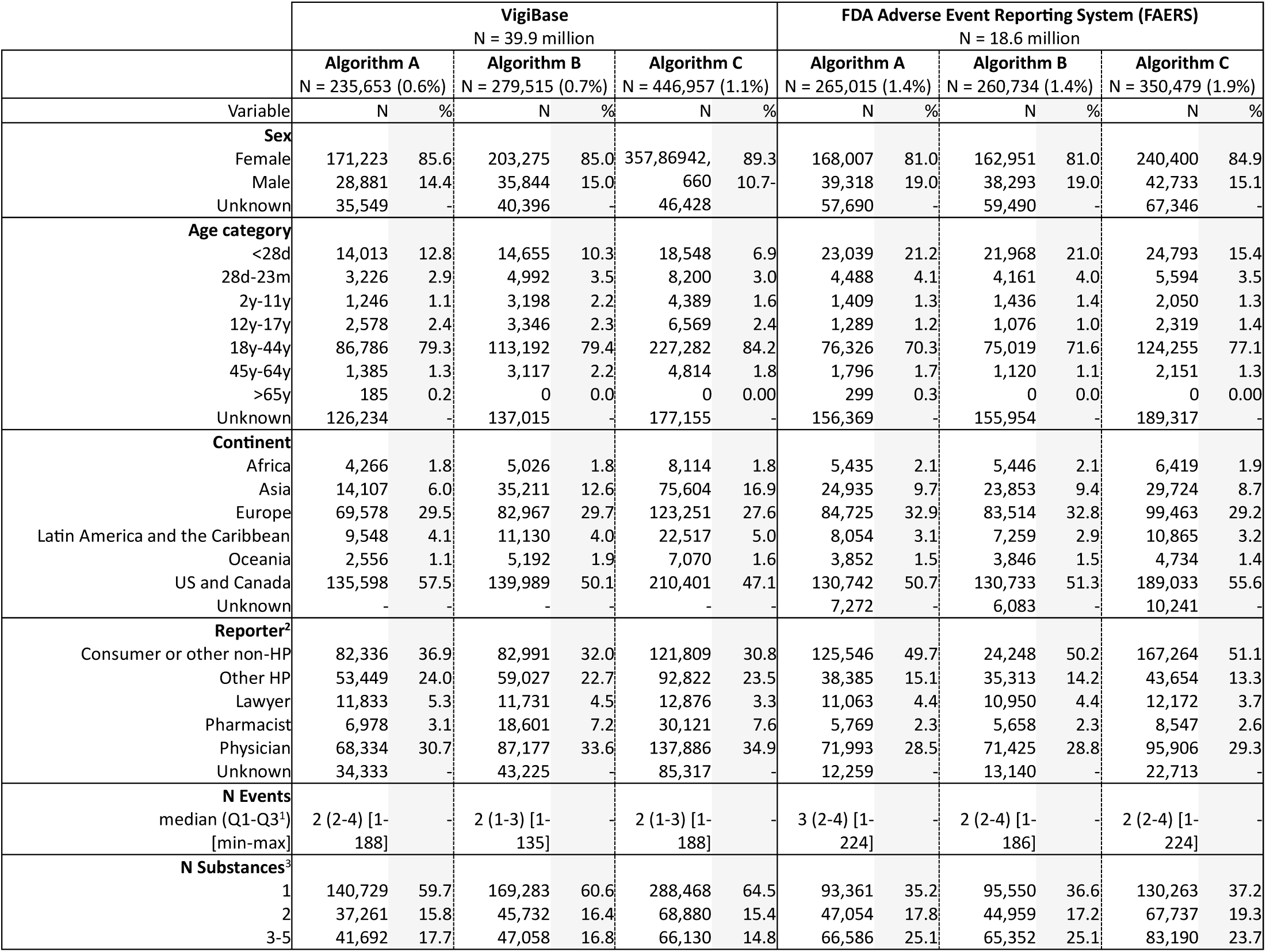
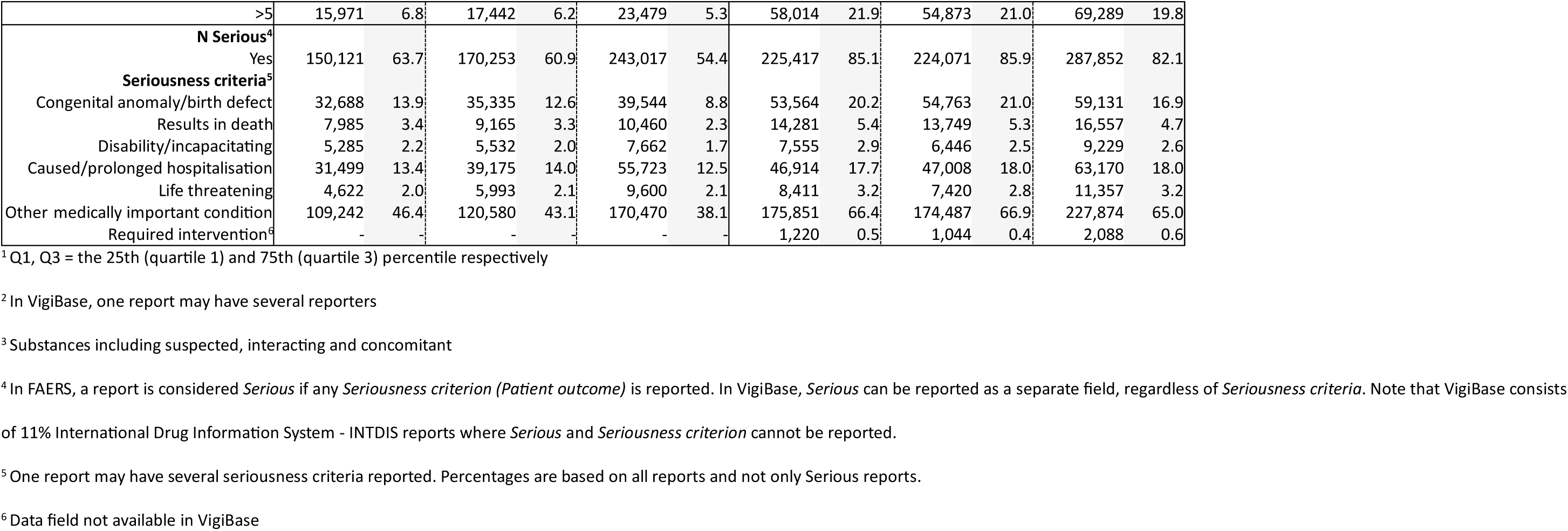
Main characteristics of reports retrieved by the three algorithms in FDA adverse event reporting system, FAERS, and the WHO global database of adverse event reports, VigiBase. HP = Health professional.

### 4.3 Comparison of algorithm rules influencing retrieval

The distributions of rules met for different algorithmic overlaps are shown in **Fig. 3** (VigiBase) and **Fig. 4** (FAERS).

**Fig. 3.**
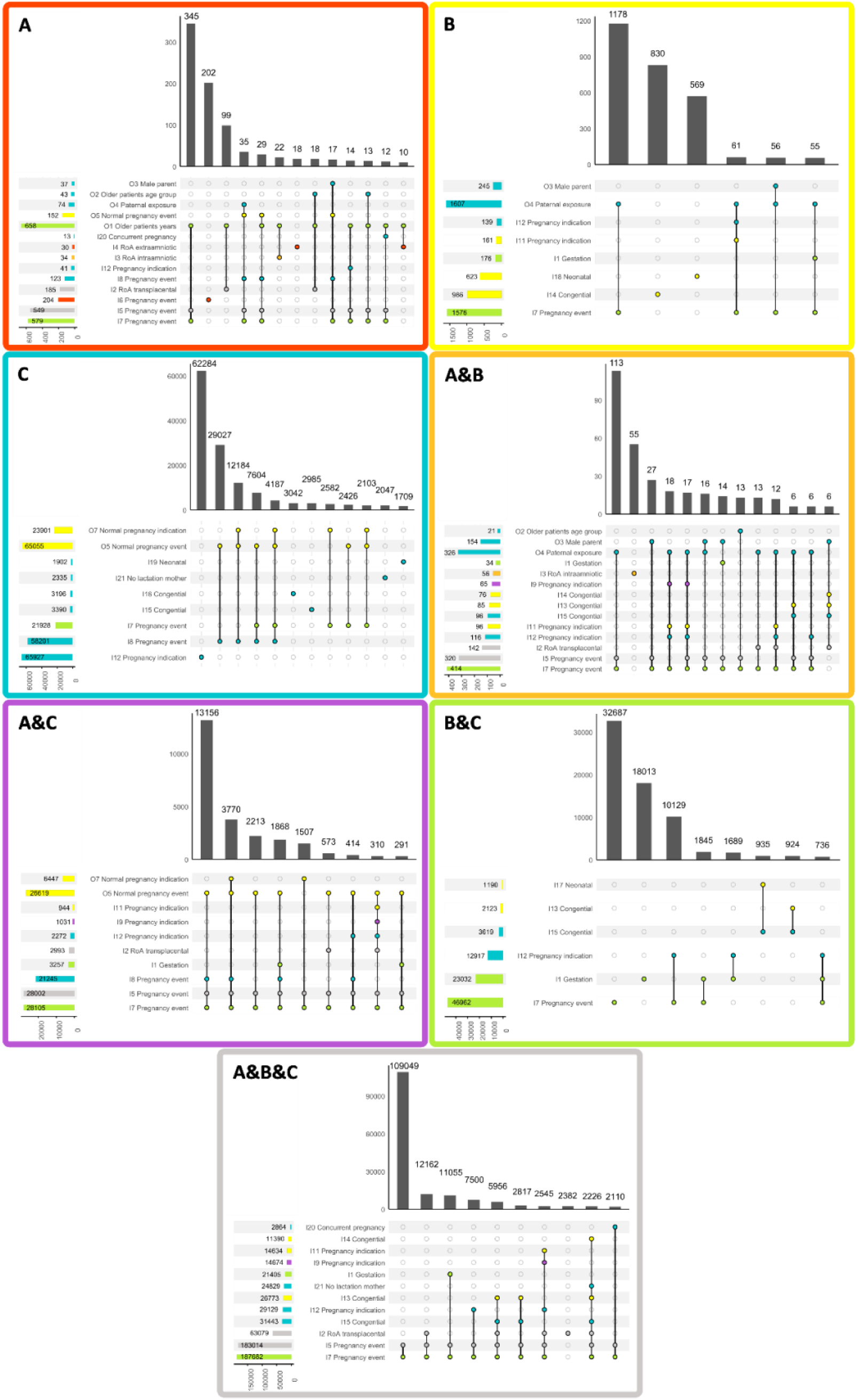
Upset plots showing the distribution of rules met when flagged by the different algorithms in *<u>VigiBase</u>*, flagged by only A, only B, only C, A and B but not C, A and C but not B, B and C but not A, and flagged by all three algorithms A, B and C. Only combinations with at least 1% of the reports of each group are shown.

**Fig. 4.**
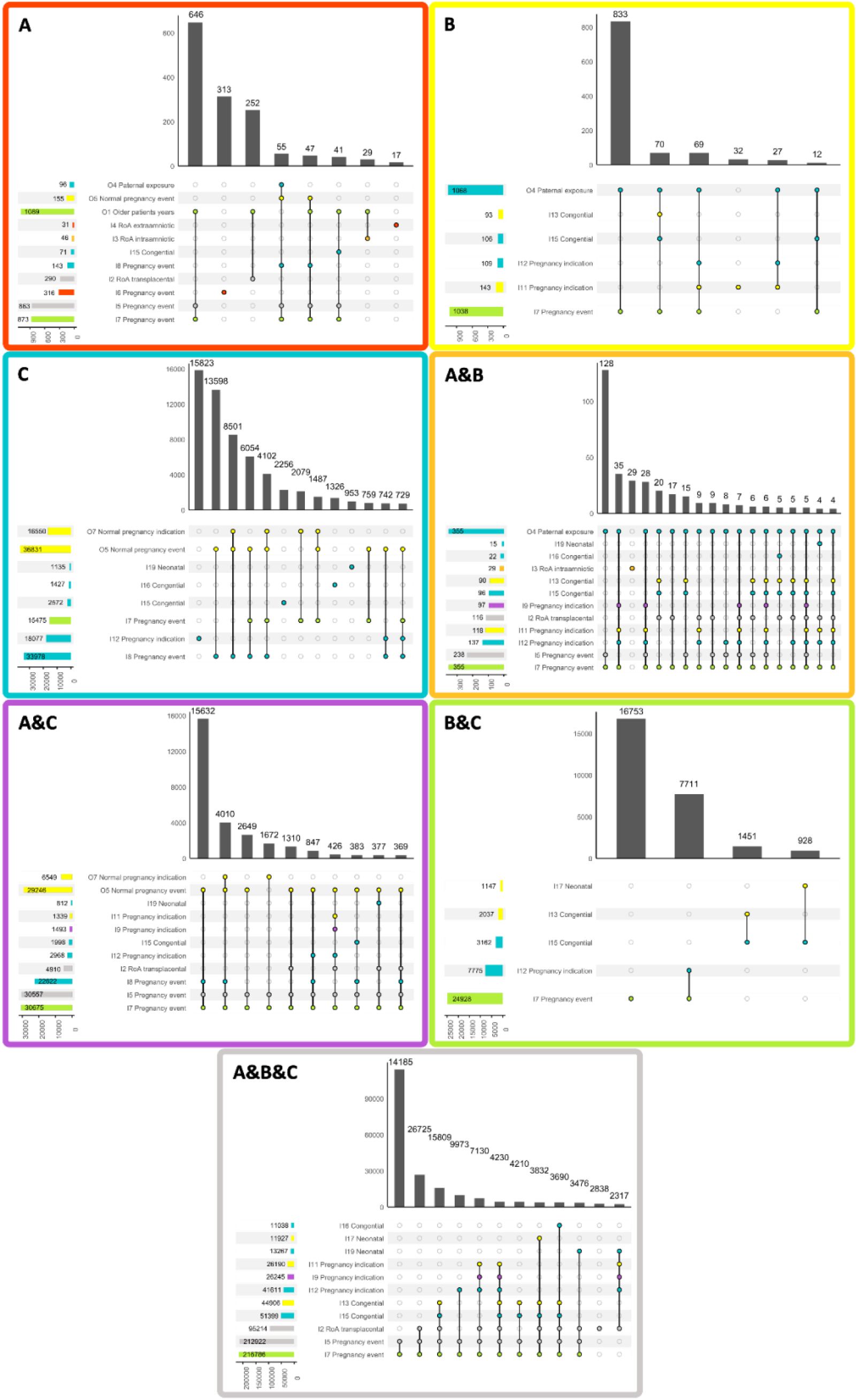
Upset plots showing the distribution of rules met when flagged by the different algorithms in *<u>FAERS</u>*, flagged by only A, only B, only C, A and B but not C, A and C but not B, B and C but not A, and flagged by all three algorithms A, B and C. Only combinations with at least 1% of the reports of each group are shown.

#### 4.3.1 Reports retrieved by all three algorithms

Among the 205285;231517 reports flagged by all the algorithms (in VigiBase;in FAERS), 183014;212922 (89%;92%) included a pregnancy-related exposure PT among the adverse events (I5; subset of I7 terms). The second most common criterion met was transplacental route of administration (I2), reported in total in 63079;95214 (31%;41%) retrieved reports, and as the only criterion met in 2382;2838 (1%;1%) reports.

#### 4.3.2 Reports retrieved only by Algorithm A

Among the 994;1528 reports flagged only by Algorithm A, 549;863 (55%;56%) included an adverse event PT concerning pregnancy-related exposure (I5). Reports were excluded by Algorithms B and C due to different rules, mainly patient age above 50 years (O1) (658;1089, 66%;71%). Algorithm A exclusively flagged 204;316 (21%;20%) reports due to reporting ‘maternal exposure during delivery’^4^ among the adverse events (I6), and 30;31 (3%;2%) due to reporting ‘extraamniotic’ as a route of administration (I4).

#### 4.3.3 Reports retrieved only by Algorithm B

Among the 3248;1100 reports retrieved only by Algorithm B, 1576;1038 (49%;94%) had an adverse event PT concerning foetal disorder, pregnancy or its termination (I7). This rule is in common with Algorithm C, but reports were excluded by this algorithm mainly because they referred to a paternal exposure (O4). In the VigiBase dataset specifically, 986 (30%) and 623 (19%) reports were exclusively retrieved by Algorithm B because they recorded a congenital or neonatal event, together with information about the parent of the patient (i.e., parent-child reports, which information is only available in the VigiBase dataset) (I14, I18).

#### 4.3.4 Reports retrieved only by Algorithm C

Among the 142324;59643 reports retrieved only by Algorithm C, 65927;18077 (46%;30%) were included due to an indication relating to foetal disorder, pregnancy or its termination, or normal pregnancy (I12), a rule that is exclusive to Algorithm C. All the 58201;33978 (41%;57%) reports with an adverse event referring to a normal pregnancy (I8) were excluded by Algorithm B due to excluding reports with normal pregnancy terms (O5). Of 21928;15475 (15%;26%) reports with an adverse event concerning foetal disorders, pregnancy or its termination (I7; rule for both B and C), 11924;7739 (8%;13%) reports were excluded by Algorithm B due to adverse event referring to a normal pregnancy (O5), 2697;2110 (2%;4%) due to menstrual disorders or contraceptive methods among the indications (O7), and 7307; 5626 (5%;9%) due to both O5 and O7. Other reports were exclusively included by Algorithm C due to seriousness criteria congenital anomaly/birth defect (I15), a congenital event in a child of less than 2 years (I16), a female parent without specifying exposure through lactation (I21) (parent information only available in VigiBase), or a neonatal event in a child of less than 8 days (I19).

#### 4.3.5 Reports retrieved only by Algorithm A and B

Algorithms A and B flagged 504;384 reports, that were not flagged by Algorithm C. The majority of these, 326;355 (65%;92%) were excluded by C because of an adverse event referring to paternal exposure (O4). Specifically, in VigiBase, 154 (31%) reports were excluded by C due to parental information specifying male sex (O3) and 21 (4%) reports with age group reported as elderly (O2) (parental and age group information only available in VigiBase^5^). In addition, 56;29 (11%;8%) reports were selectively included by A and B because of intra-amniotic route of administration (I3).

#### 4.3.6 Reports retrieved only by Algorithm A and C

Among the 28870;31586 reports retrieved only by A and C, 26619;29246 (92%;93%) were excluded by B due to adverse event PTs relating to normal pregnancy (O5), 6447;6549 (22%;21%) or due to indications relating to menstrual disorders or contraceptive methods (O7).

#### 4.3.7 Reports retrieved only by Algorithm B and C

Among the 70478;27733 reports retrieved only by B and C, 46962;24928 (67%;90%) recorded the B and C rule of adverse events relating to foetal disorder, pregnancy or its termination (I7). Specifically, in VigiBase, 23032 (33%) of the reports retrieved recorded information about gestation (I1). Some additional reports recorded a congenital anomaly among the seriousness criteria (I15; Algorithm C) together with either a congenital or neonatal reaction (I13, I17; Algorithm B).

## 5. Discussion

### 5.1 Main Findings

This study highlights similarities and differences in the construction and outputs of three recently developed rule-based algorithms designed to retrieve reports related to pregnancy exposures in adverse event reporting databases. While all three algorithms aim to retrieve reports concerning either the pregnant person or the foetus/prenatally exposed child (without distinguishing between the two), their intended scopes differ, which explains the main differences in output.

Algorithm A aims to retrieve pregnancy reports with high PPV, although this measure has not been formally evaluated. It neither actively includes nor excludes paternal exposures and has no restrictions on maternal age. This is the most uncomplicated algorithm to implement and interpret. Algorithm B only targets reports with adverse events and actively retrieves paternal exposures in addition. Algorithm C adopts a broader scope, including normal and unintended pregnancies, while excluding paternal exposures.

Besides differences in scope, all three algorithms are rule-based and use structured fields only. They all have in common that they use pregnancy exposure terms and transplacental route of administration for ruling in reports, however, the exact exposure terms somewhat differ and two of the algorithms (A and B) use additional routes of administration for inclusion. Algorithm A operates without additional rules while, Algorithms B and C^6^ use strict exclusion criteria. While some exclusion rules were helpful in ruling out reports that are not likely to be related to pregnancy, there is also a risk to miss relevant reports with other eligible information at later stages. For example, B and C rule out reports based on high age as a mean to reduce false positives, although this may result in missing true pregnancy cases. Another example is B excluding reports indicating a normal pregnancy, despite potentially co-reporting other events indicating an adverse event, and the exclusion of paternal exposures by Algorithm C (only when maternal exposure is not co-reported, aiming to retain reports with exposures through both parents). Algorithm A, being more stringent in its criteria for ruling in reports, may rely less on such exclusion rules.

Both Algorithm B and C rule in reports with events referring to congenital anomalies or neonatal disorders, although these categories may reflect historical or neonatal exposures. To mitigate the risk of false positives, Algorithm B additionally requires congenital anomaly/birth defect to be reported as a seriousness criterion, or the report to be a parent-child report. Algorithm C, on the other hand, restricts the rule based on patient age (as already including reports solely based on the seriousness criterion or the presence of a female parent). As a result of these different design choices, reports with a congenital anomaly/birth defect seriousness criterion but no congenital term among the events would be flagged by Algorithm C but not by B. Among these, we find both reports lacking specific congenital event terms due to incomplete reporting (highlighting the importance of better information capture) and reports where seriousness may be miscoded [16].

The versions of Algorithm A intending to be more inclusive are more alike Algorithm B and C in their setup of rules. Some differences in the design then stem from differences in the structure of the database for which the algorithm was developed. For example, Algorithms B and C use information about gestation period, parent of the patient, and concurrent condition (part of medical history section; C only), which was not available during the development of Algorithm A. Moreover, the availability of additional information in VigiBase compared to FAERS explain some of the differences in reports retrieved selectively by only one or two of these algorithms.

A limitation of the current implementations of Algorithms B and C is that some rules rely solely on the presence of a populated field, without validating its content. As a result, coding errors may lead to misclassification.

Algorithm C flagged the highest number of reports as pregnancy-related, particularly when applied to VigiBase. Most reports flagged by Algorithms A and B were also retrieved by C, suggesting higher sensitivity (while of course dependant on the scope in mind). However, this study did not assess PPV, so it remains unclear whether this potentially improved sensitivity comes at the expense of PPV. Prior evaluations reported PPV rates of 90% and 92% for Algorithms B and C, respectively, within their intended scope ^7^ [15, 16]. Furthermore, additional reports flagged by C may regard normal and unintended pregnancies which is outside the scope of B. This, alongside with Algorithm B’s inclusion of paternal exposures, might explain the higher proportion of reproductive-age females and the lower proportion of neonatal reports (despite higher absolute numbers) flagged by C.

Notably, all algorithms flagged a higher proportion of pregnancy-related reports in FAERS compared to VigiBase. This may reflect the greater proportion of serious reports in FAERS, as pregnancy-related reports are more frequently reported as serious. In fact, unlike VigiBase, FAERS receives only serious reports from non-US countries. Additionally, around 11% of VigiBase reports were submitted in the older International Drug Information System – INTDIS format, which lacked several structured fields, making it more challenging for the algorithms to classify them as pregnancy-related.

### 5.2 Strengths and Limitations

A main strength of this study is the collaborative effort among the developers of all three algorithms. This ensured that each algorithm was implemented and interpreted in line with its original design, and that the conclusions drawn are agreed from all perspectives.

Another strength lies in the application of the algorithms to two major adverse event databases, VigiBase and FAERS, which differ in structure and available data fields, offering a broader perspective on algorithm performance. Algorithm B was originally developed for EudraVigilance (based on the ICH-E2B(R3) format), which was not included due to feasibility constraints. This limits the observation of Algorithm B in its native context.

Another limitation is that we did not systematically assess the accuracy of the algorithms in flagging true pregnancy-related reports. Instead, focus was on characterising the relative outputs across databases, and we refer to prior PPV evaluations of Algorithms B and C. Reports missed by the algorithms were also not examined, and we can only relate the sensitivity to the prior evaluation of Algorithm C.

### 5.3 Future Directions

Each algorithm offers valuable insights that lay the foundation for further improvements. Hybrid approaches that combine strengths from each algorithm could potentially support the development of more flexible and context-driven algorithms. While the algorithms provide structured approaches to flag pregnancy-related reports, their optimal use in signal detection and exposure characterisation remains to be explored.

In parallel to algorithm refinements, it is equally important to improve data quality in order to enhance the retrieval and utilisation of pregnancy data. Reports should clearly indicate pregnancy by reporting an exposure term among the events [21], and include key details such as discriminating between maternal and paternal exposure, indicating the precise timing of exposure in relation to gestational age, describing comorbidities and medical history. Consistency in describing the report subject, avoiding inconsistencies like patient age referring to the pregnant person while adverse outcomes to the child, is essential [16]. Harmonisation, coding guidelines and regular training can enhance the capture of pregnancy-related data. Underreporting remains a significant issue, with many adverse events occurring in pregnancy never documented [22, 23], and must also be addressed.

As already discussed, there is currently no definitive indicator to confirm whether a report truly concerns exposure related to pregnancy or not. A report may be miscoded as pregnancy-related, or may concern a pregnant person without any indicators of pregnancy in the report. Therefore, to differentiate between pregnancy-related and non-pregnancy-related reports, we must first operationalise the construct of pregnancy itself. Each algorithm represents a different operational definition, with different sets of rules. Going forward, two paths are possible. One is to establish a common standardised algorithm, promoting consistency, comparability and replicability across studies. The other is to embrace the diversity of approaches, equipping researchers with tools to understand and select or tailor algorithms for their specific needs. Recognising the distinct features and scopes of the algorithms, we chose to retain this pluralism, promoting transparency and informed decision-making. Additionally, improving information capture at the point of the collection, ensuring that new reports include a reliable flag for pregnancy exposure, would be the ideal scenario.

Ultimately, the three algorithms represent different conceptualisations of pregnancy-related reports with some overlaps. Understanding not just what each algorithm retrieves, but also how and why it does so, is critical for advancing pregnancy-specific pharmacovigilance.

## 6. Conclusion

While there are significant commonalities among the compared algorithms used for flagging reports related to exposure during pregnancy, they partly differ in scopes, utilised data, and specific rules. These commonalities and differences are reflected in the number and type of reports each algorithm retrieves and the overlap across them.

By characterising the design and outputs of each algorithm, this study provides pharmacovigilance professionals with the insights needed to select the most appropriate approach for their specific needs and informs further developments of the algorithms. Rather than advocating for a single standard, we suggest a pluralistic and transparent way forward that supports informed decision-making based on context and purpose.

Looking ahead, a crucial aspect is improving data capture at the point of collection. Ensuring that future reports include reliable indicators of pregnancy exposure would reduce reliance on retrospective algorithms and enhance pregnancy-related pharmacovigilance.

## Declarations

### Funding

The authors received no financial support for the research, authorship, and/or publication of this article. Nhung Trinh reports to receive internationalization funding from UiO Life Science initiative to organize the workshop related to this work in Oslo in January 2025.

### Conflicts of interest

All authors declare no conflicts of interest.

### Ethics approval

Not applicable. This study did not use personal data.

### Consent to participate

Not applicable.

### Consent for publication

Not applicable.

### Code availability

The code used for this study is available at https://github.com/Uppsala-Monitoring-Centre/PVgravID.

### Author contributions

All authors contributed to the study conception and design. Material preparation and data collection were performed by Michele Fusaroli, Valentina Giunchi, Levente K-Pápai, Takamasa Sakai and Sara Hedfors Vidlin. All authors contributed to the analysis. The first draft of the manuscript was written by Nhung TH Trinh, Elena Rocca, Lovisa Sandberg, Michele Fusaroli, Levente K-Pápai, Valentina Giunchi and Sara Hedfors Vidlin, and all authors commented on previous versions of the manuscript. All authors read and approved the final manuscript.

## Data Availability

The data that support the findings of this study are partly publicly available. The FAERS database is publicly available in raw form at https://fis.fda.gov/extensions/FPD-QDE-FAERS/FPD-QDE-FAERS.html, and in a clean version through DiAna. An open access toolkit for DIsproportionality ANAlysis and other pharmacovigilance investigations in the FAERS (https://github.com/fusarolimichele/DiAna_package). Access to VigiBase is restricted based on the conditions for access within the WHO Programme for International Drug Monitoring. Subject to these conditions, data is available from the authors on reasonable request. For further inquiries, please contact Uppsala Monitoring Centre via https://who-umc.org/contact-information/.

The VigiBase pregnancy algorithm (Algorithm C) is implemented in VigiLyze, the tool for searching and analysing VigiBase data, which is available to national pharmacovigilance centres as members of the WHO Programme for International Drug Monitoring. VigiBase data, including output of the VigiBase pregnancy algorithm, is also available through VigiBase Custom Searches.

## Acknowledgements

The authors would like to express our deepest gratitude to Professor Angela Lupattelli, whose insight, dedication, and passion were instrumental in the conception, design, and analysis of this study. Although she sadly passed away during the course of the work, her influence is present throughout this study. We remember her with great respect and appreciation.

The views expressed in this article are the personal views of the author(s) and may not be understood or quoted as being made on behalf of or reflecting the position of the regulatory agency/agencies or organisations with which the author(s) is/are employed/affiliated.

The views expressed are those of the authors and not necessarily those of Uppsala Monitoring Centre, University of Bologna,, Metropolitan University of Oslo, Meijo University, or University of Oslo.

The Uppsala Monitoring Centre authors are indebted to the national centres which make up the WHO Programme for International Drug Monitoring and contribute reports to VigiBase. However, the opinions and conclusions of this study are not necessarily those of the various centres nor of the WHO. MedDRA® trademark is registered by ICH.

## Footnotes

1 The MedDRA® Medical Dictionary for Regulatory Activities terminology is the international medical terminology developed under the auspices of the International Council for Harmonisation of Technical Requirements for Pharmaceuticals for Human Use (ICH).

2 Note that this algorithm has not been developed by or in collaboration with US FDA.

3 In this study, we have aligned the FAERS field *Patient outcome* (comprising *Death*, *Life-threatening*, *Hospitalization*, *Disability*, *Congenital anomaly*, *Other serious outcome*, *Required intervention*) with the ICH-E2B field *Seriousness criteria* (comprising *Results in death*, *Life threatening*, *Caused/prolonged hospitalisation*, *Disabling/incapacitating*, *Congenital anomaly/birth defect*, *Other medically important condition*). We will refer to this as seriousness criteria everywhere. In FAERS, a case is considered serious only if it has at least one *Patient outcome* reported. In VigiBase, a case can be reported as serious even without any seriousness criteria.

4 ‘Maternal exposure during delivery’ is not included in the narrow scope of ‘Pregnancy and neonatal topics’ SMQ, which is the basis for reaction terms included by Algorithm B and C.

5 Reported age group information exist in the original extract of FAERS but is not available in the data extract from DiAna. Age groups in DiAna are recalculated from onset age and therefor completely redundant, considering rule O1.

6 and more inclusive versions of algorithm A

7 For algorithm C also sensitivity was evaluated.

## Notes

### Competing Interest Statement

The authors have declared no competing interest.

### Author Declarations

Ethical board review was not required for this study, as this study did not use personal data. Data was used according to Data Access Conditions from the VigiBase WHO database of adverse event reports and the openly available FDA Adverse Event Reporting System database.

