## Supplementary Material for "Comparing Existing Algorithms for Retrieving Pregnancy-related Adverse Event Reports"

### Electronic Supplementary Material

**Article title:** Exploring and Comparing Existing Algorithms for Flagging Pregnancy-related Adverse Event Reports

**Journal:** Drug Safety

### S1: Algorithm rules

**Table S1** The algorithm rules and corresponding fields and values used.

PT = MedDRA Preferred Term, LLT = MedDRA Lowest Level Term, HLT = MedDRA High Level Term, HLGT = MedDRA High Level Term = High Level Group Term, SMQ = Standardized MedDRA Queries (all use of SMQ referring to level 2 sub-SMQs in the 'Pregnancy and neonatal topics (SMQ)', narrow scope)

| Algorithm rule | Action | Applicable to algorithm | | | Field | | Data |
|---|---|---|---|---|---|---|---|
| | | A | B | C | | | |
| O1 | Rule out | | X | X | Age at time of onset of reaction/event | > | 50 years |
| O2 | Rule out | | | X | Patient age group | = | Elderly |
| O3 | Rule out | | | X | Sex of parent | = | Male |
| O4 | Rule out | | | X | Reaction(s)/ event(s) | in | Paternal exposure term α:<br>*Maternal exposure via partner during pregnancy (PT)* [1,2]<br>*Paternal exposure before pregnancy (PT)* [1,2]<br>*Paternal exposure during pregnancy (PT)* [1,2]<br>*Paternal exposure timing unspecified (PT)* [1,2]<br>Paternal exposure term β:<br>*Paternal drugs affecting foetus (PT)* [3]<br>Paternal exposure term δ:<br>*Exposure via father (PT)* [* 2]<br>*Pregnancy of partner (PT)* [4]<br>*Exposure via partner (PT)*<br>*Transmission of drug via semen (LLT)*<br>*Exposure via semen (LLT)* |
| | | | | | AND | | |
| | | | | | Reaction(s)/ event(s) | not in | Maternal exposure term α:<br>*Maternal exposure before pregnancy (PT)* [1,2]<br>*Maternal exposure during pregnancy (PT)* [1,2]<br>*Maternal exposure timing unspecified (PT)* [1,2]<br>Maternal exposure term β:<br>*Maternal exposure during delivery (PT)* [1]<br>Maternal exposure term γ:<br>*Maternal drugs affecting foetus (PT)* [3] |
| O5 | Rule out | | X | | Reaction(s)/ event(s) | in | *Normal pregnancy conditions and outcomes (SMQ)*<br>*Lactation related topics (incl neonatal exposure through breast milk) (SMQ)*<br>*Menstrual cycle and uterine bleeding disorders (HLGT)*<br>*Contraceptive methods female (HLT)*<br>*Ectopic pregnancy under hormonal contraception (PT)* [2]<br>*Ectopic pregnancy with contraceptive device (PT)* [2] |
| O6 | Rule out | | X | | Reaction(s)/ event(s) | | Only exposure term, without any term considering an adverse outcome |
| O7 | Rule out | | X | | Indication | in | *Menstrual cycle and uterine bleeding disorders (HLGT)*<br>*Contraceptive methods female (HLT)* |
| I1 | Rule in | | X | X | Gestation period when reaction/event was observed in the foetus | ≠ | NULL |
| | | | | | OR | | |
| | | | | | Gestation period at time of exposure | ≠ | NULL |
| I2 | Rule in | X | X | X | Route of administration | = | Transplacental |
| I3 | Rule in | X | X | | Route of administration | = | Intraamnoitic |

| | | | | | | | |
|---|---|---|---|---|---|---|---|
| I4 | Rule in | X | | | Route of administration | = | Extraamnoitic |
| I5 | Rule in | X | X | X | Reaction(s)/ event(s) | in | Maternal exposure term α:<br>*Maternal exposure before pregnancy (PT)* [1,2]<br>*Maternal exposure during pregnancy (PT)* [1,2]<br>*Maternal exposure timing unspecified (PT)* [1,2]<br>Maternal exposure term γ:<br>*Maternal drugs affecting foetus (PT)* [3]<br>General exposure term α:<br>*Drug exposure before pregnancy (PT)* [1,2]<br>*Exposure during pregnancy (PT)* [1,2]<br>*Foetal exposure during delivery (PT)* [1,2]<br>*Foetal exposure during pregnancy (PT)* [1,2]<br>*Foetal exposure timing unspecified (PT)* [1,2] |
| I6 | Rule in | X | | | Reaction(s)/ event(s) | in | Maternal exposure term β:<br>*Maternal exposure during delivery (PT)* [1] |
| I7 | Rule in | | X | X | Reaction(s)/ event(s) | in | *Pregnancy, labour and delivery complications and risk factors (excl abortions and stillbirth) (SMQ)*<br>*Foetal disorders (SMQ)*<br>*Termination of pregnancy and risk of abortion (SMQ)* |
| I8 | Rule in | | | X | Reaction(s)/ event(s) | in | *Normal pregnancy conditions and outcomes (SMQ)* |
| I9 | Rule in | X | | X | Indication | in | Maternal exposure term α:<br>*Maternal exposure before pregnancy (PT)* [1,2]<br>*Maternal exposure during pregnancy (PT)* [1,2]<br>*Maternal exposure timing unspecified (PT)* [1,2]<br>Maternal exposure term γ:<br>*Maternal drugs affecting foetus (PT)* [3]<br>General exposure term α:<br>*Drug exposure before pregnancy (PT)* [1,2]<br>*Exposure during pregnancy (PT)* [1,2]<br>*Foetal exposure during delivery (PT)* [1,2]<br>*Foetal exposure during pregnancy (PT)* [1,2]<br>*Foetal exposure timing unspecified (PT)* [1,2] |
| I10 | Rule in | X | | | Indication | | Maternal exposure term β:<br>*Maternal exposure during delivery (PT)* [1] |
| I11 | Rule in | | X | | Indication | in | Maternal exposure term α:<br>*Maternal exposure before pregnancy (PT)* [1,2]<br>*Maternal exposure during pregnancy (PT)* [1,2]<br>*Maternal exposure timing unspecified (PT)* [1,2]<br>Maternal exposure term β:<br>*Maternal exposure during delivery (PT)* [1]<br>General exposure term α:<br>*Drug exposure before pregnancy (PT)* [1,2]<br>*Exposure during pregnancy (PT)* [1,2]<br>*Foetal exposure during delivery (PT)* [1,2]<br>*Foetal exposure during pregnancy (PT)* [1,2]<br>General exposure term β:<br>*Exposed to maternal infection in utero (PT)* ** [1,2]<br>Paternal exposure term α:<br>*Maternal exposure via partner during pregnancy (PT)* [1,2]<br>*Paternal exposure before pregnancy (PT)* [1,2]<br>*Paternal exposure during pregnancy (PT)* [1,2]<br>*Paternal exposure timing unspecified (PT)* [1,2]<br>Paternal exposure term γ:<br>*Foetal exposure via father (PT)* ** [1,2] |
| | | | | | AND | | |
| | | | | | Indication | not in | *Lactation related topics (incl neonatal exposure through breast milk) (SMQ)* |
| I12 | Rule in | | | X | Indication | in | *Pregnancy, labour and delivery complications and risk factors (excl abortions and stillbirth) (SMQ)*<br>*Foetal disorders (SMQ)*<br>*Termination of pregnancy and risk of abortion (SMQ)*<br>*Normal pregnancy conditions and outcomes (SMQ)* |

| | | | | | | | |
|---|---|---|---|---|---|---|---|
| I13 | Rule in | | X | | Reaction(s)/ event(s) | in | *Congenital, familial and genetic disorders (SMQ)* |
| | | | | | AND | | |
| | | | | | Seriousness criterion: Congenital anomaly/birth defect | = | True |
| I14 | Rule in | | X | | Reaction(s)/ event(s) | in | *Congenital, familial and genetic disorders (SMQ)* |
| | | | | | AND | | |
| | | | | | Parent-child/foetus report | ≠ | NULL |
| I15 | Rule in | | | X | Seriousness criterion: Congenital anomaly/birth defect | = | True |
| I16 | Rule in | | | X | Reaction(s)/ event(s) | in | *Congenital, familial and genetic disorders (SMQ)* |
| | | | | | AND | | |
| | | | | | Age at time of onset of reaction/event | < | 2 years |
| I17 | Rule in | | X | | Reaction(s)/ event(s) | in | *Neonatal disorders (SMQ)* |
| | | | | | AND | | |
| | | | | | Seriousness criterion: Congenital anomaly/birth defect | = | True |
| I18 | Rule in | | X | | Reaction(s)/ event(s) | in | *Neonatal disorders (SMQ)* |
| | | | | | AND | | |
| | | | | | Parent-child/foetus report | ≠ | NULL |
| I19 | Rule in | | | X | Reaction(s)/ event(s) | in | *Neonatal disorders (SMQ)* |
| | | | | | AND | | |
| | | | | | Age at time of onset of reaction/event | < | 8 days |
| I20 | Rule in | | | X | Medical history | in | *Pregnancy (PT) except LLTs specifying "birth" or "delivered"*<br>*Multiple pregnancy (PT) except LLTs specifying "birth" or "delivered"*<br>*Twin pregnancy (PT) except LLTs specifying "birth" or "delivered"* |
| | | | | | AND | | |
| | | | | | Continuing | = | True |
| I21 | Rule in | | | X | Sex of parent | = | Female |
| | | | | | AND | | |
| | | | | | Reaction(s)/ event(s) | not in | *Lactation related topics (incl neonatal exposure through breast milk) (SMQ)* |
| | | | | | AND | | |
| | | | | | Route of administration | ≠ | Transmammary |

[*] PT available in MedDRA 26.1, at the time of development of Algorithm C, but replaced with *Foetal exposure via father* in ≥ 27.0
[**] PT not available until MedDRA ≥ 27.0, after development of Algorithm C

[1] PT is in *Exposures associated with pregnancy, delivery and lactation (HLT)*

[2] PT is in *Pregnancy, labour and delivery complications and risk factors (excl abortions and stillbirth) (SMQ)*
[3] PT is in *Foetal disorders (SMQ)*
[4] PT is in *Normal pregnancy conditions and outcomes (SMQ)*

### S2: Original implementation flowcharts

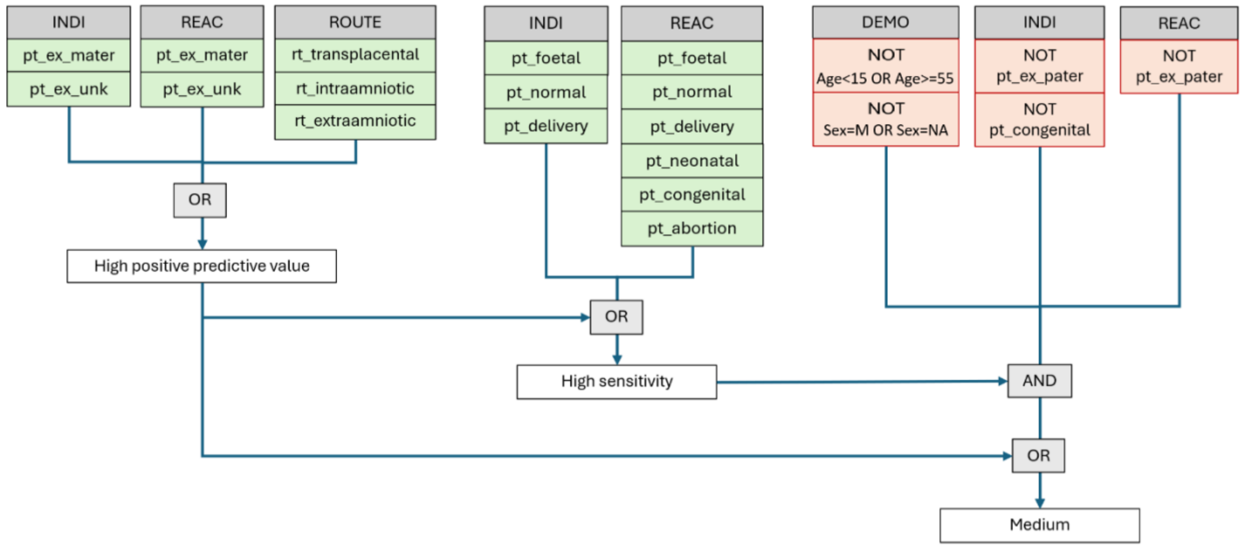


**Fig. S2:1** Sakai et al: High positive predictive value version (Algorithm A), Medium version and High sensitivity version

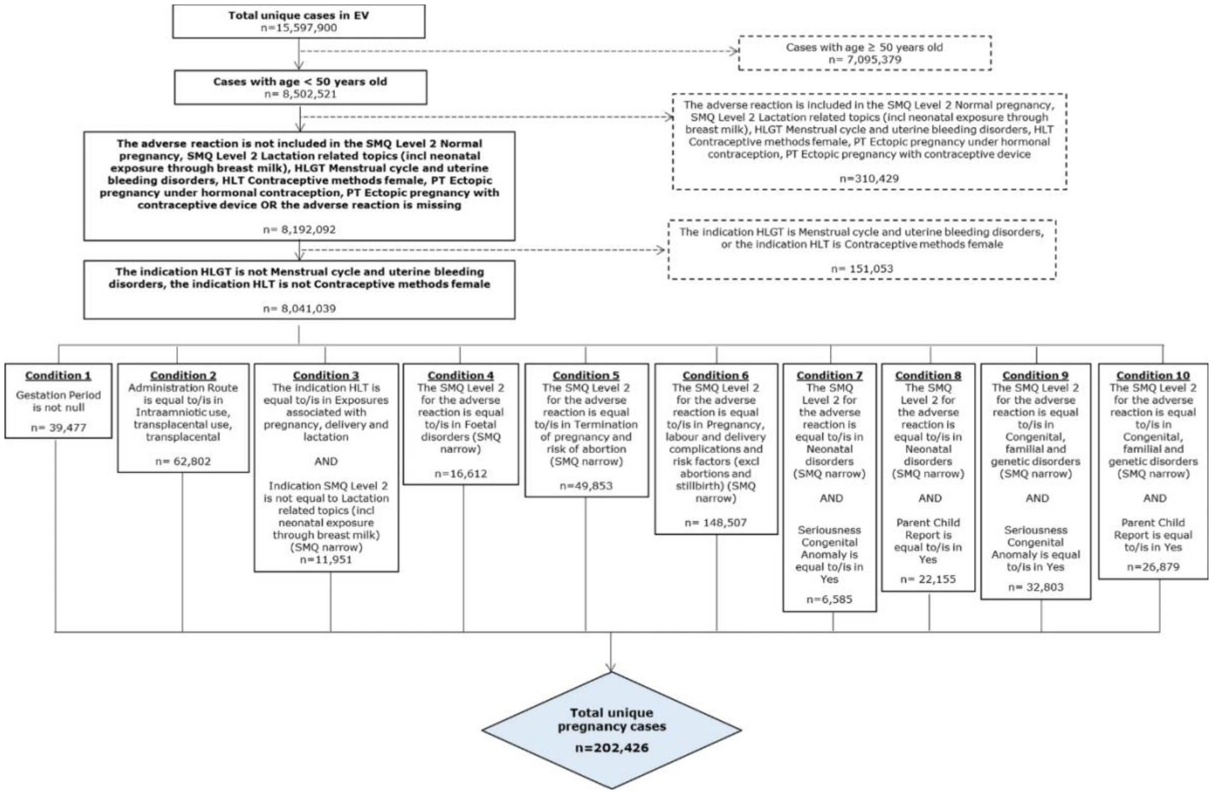


**Fig. S2:2** Zaccaria et al: Algorithm B as presented in **Zaccaria C, Piccolo L, Gordillo-Marañón M, et al (2024) Identification of Pregnancy Adverse Drug Reactions in Pharmacovigilance Reporting Systems: A Novel Algorithm Developed in EudraVigilance. Drug Saf 47:1127–1136. https://doi.org/10.1007/s40264-024-01448-y**

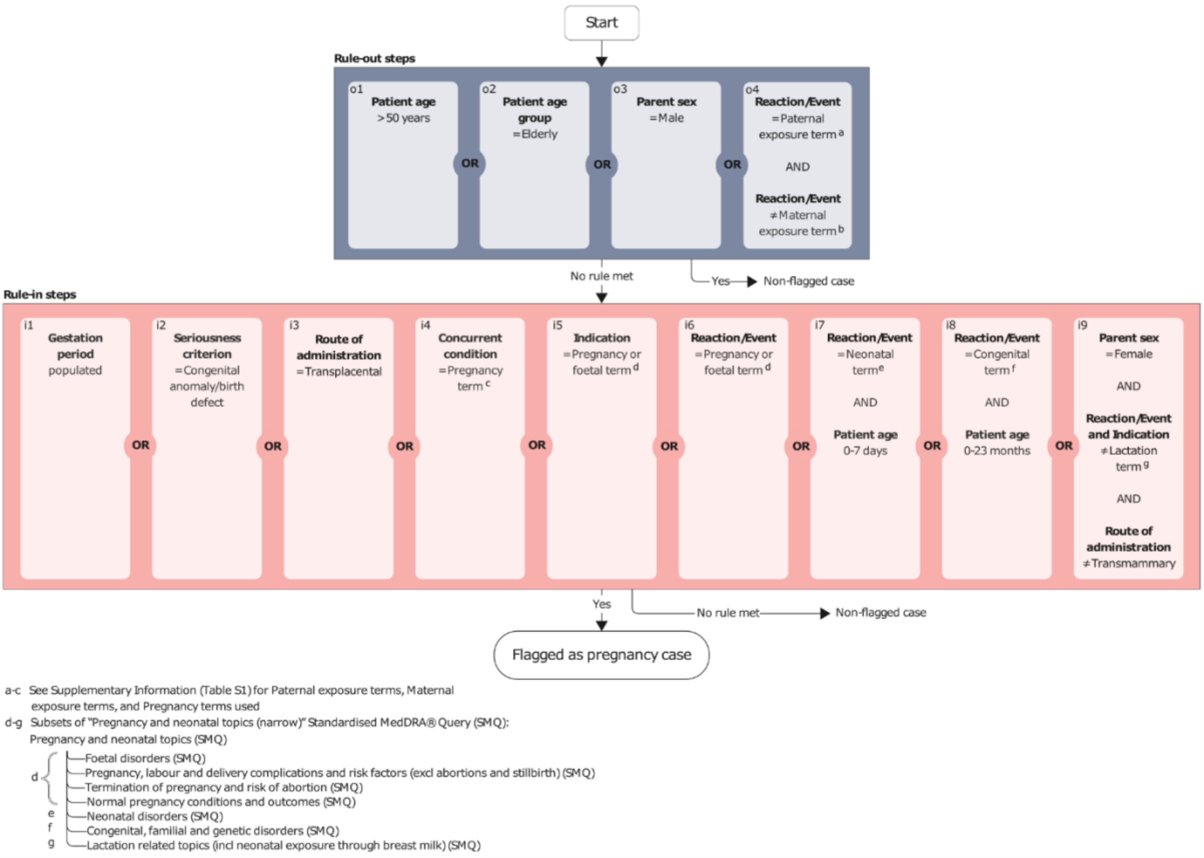


**Fig. S2:3** VigiBase pregnancy algorithm described in Sandberg et al: Algorithm C as presented in **Sandberg L, Vidlin SH, K-Pápai L, et al (2025) Uncovering Pregnancy Exposures in Pharmacovigilance Case Report Databases: A Comprehensive Evaluation of the VigiBase Pregnancy Algorithm. Drug Saf. https://doi.org/10.1007/s40264-025-01559-0**

#### S3: Result figures for Sakai et al Medium version and High sensitivity version

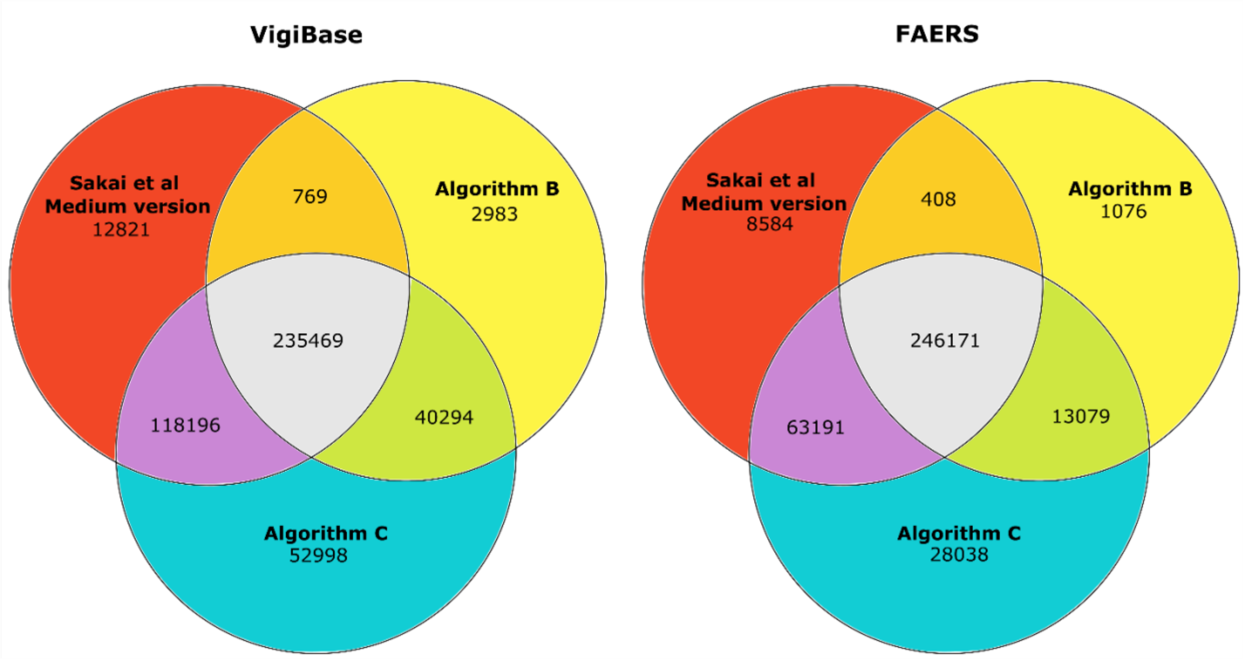


**Fig. S3:1** Venn diagrams showing the overlap of the pregnancy-related reports retrieved by the algorithms Sakai et al Medium version, Algorithm B and Algorithm C, in VigiBase and FAERS, respectively. Note that the diagrams are non-proportional.

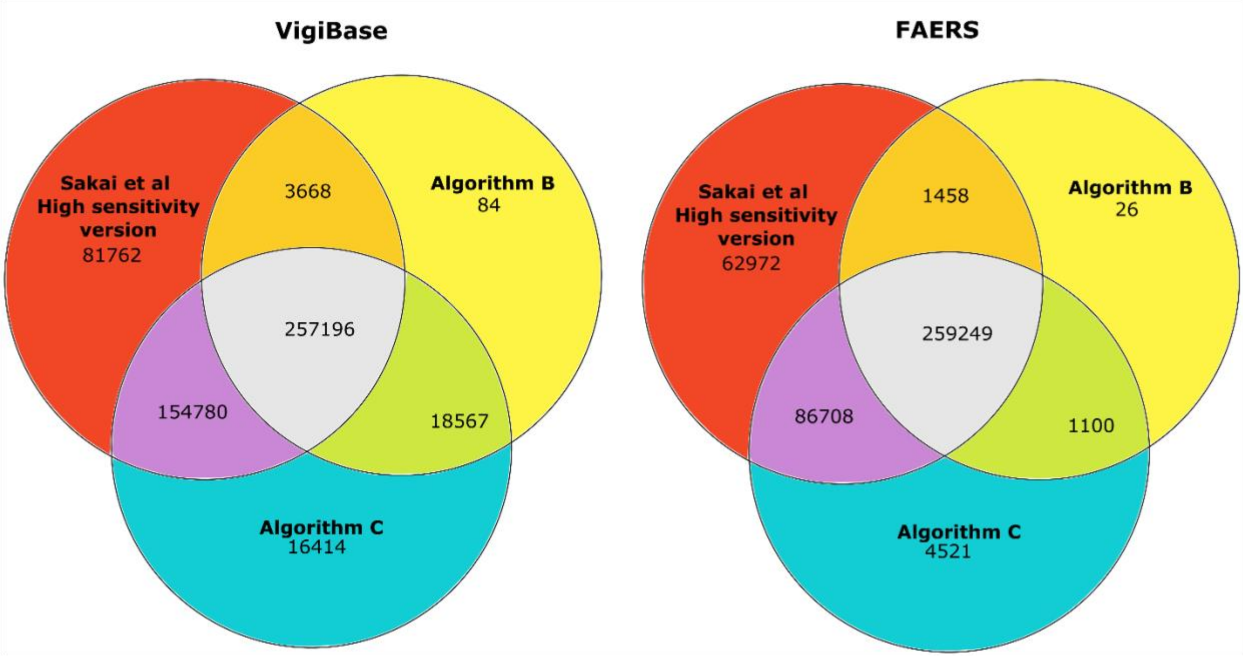


**Fig. S3:2** Venn diagrams showing the overlap of the pregnancy-related reports retrieved by the algorithms Sakai et al High sensitivity version, Algorithm B and Algorithm C, in VigiBase and FAERS, respectively. Note that the diagrams are non-proportional.
